# The mode effect of web-based surveying on the 2018 HRS measure of cognitive functioning

**DOI:** 10.1101/2022.11.03.22281879

**Authors:** Benjamin W. Domingue, Ryan McCammon, Brady T. West, Ken Langa, David Weir, Jessica Faul

**Affiliations:** Stanford Graduate School of Education; University of Michigan

## Abstract

Measuring cognition in an aging population is a public health priority. A move towards survey measurement via the web (as opposed to phone or in-person) is cost effective but challenging as it may induce bias in cognitive measures. We examine this possibility using an experiment embedded in the 2018 wave of data collection for the US Health and Retirement Study (HRS). We find evidence of an increase in scores for HRS respondents who are randomly assigned to the web-based mode of data collection in 2018. Web-based respondents score higher in 2018 than do phone-based respondents, and they show much larger gains relative to 2016 performance and subsequently larger declines in 2020. The bias in favor of web-based responding is observed across all item types, but most pronounced for the serial 7 and items on financial literacy. These results imply a larger threshold for web-based respondents in applications requiring an indicator of cognitive impairment. Implications for both use of HRS data and future survey work on cognition are discussed.

## 1 Introduction

As the population of the US and the world ages, health issues that arise as a function of this increased longevity are becoming more salient. A key such health condition is age-related decline in cognitive functioning [1]. As a result, there is an increased focus on measuring cognitive functioning in surveys that aim to understand the health and well-being of older populations. The Health and Retirement Study (HRS) [2] is designed to provide such information. While the HRS is a US-based cohort of people over 50 and their spouses, it is also the template for a broader range of global studies focused on aging [2]. The HRS undertakes a study of cognition with its respondents [3] but there are numerous challenges associated with this measurement task.

One key challenge has to do with conducting comparable measurement across different measurement modalities (e.g., phone versus face-to-face responding). From a psychometric perspective, such variation would be an example of the more general problem of measurement invariance [4]. The quality of measurement may vary as a function of survey modality; understanding this variation and its implications is essential to proper subsequent use of the resulting measures. In this particular case, there is concern that the introduction of a web-based modality for survey responding on the HRS may be “easier” than the phone-based modality given that respondents can quickly utilize computing tools (e.g., text editors, calculators) when they engage with the survey questions via the web. Compounding the challenge is that, by the nature of how HRS categorizes respondent eligibility for the cognition battery, phone- and web-based samples are not exchangeable.

This paper attempts to assess the possibility of mode effects focusing on the novel deployment of the HRS cognitive module [5] via both the web and phone in 2018. To do this, we deploy techniques from item response theory (IRT, [6]) and the differential item functioning (DIF) literatures. Our use of IRT builds on earlier work focusing on the effect of web-based administration on this survey [7] (note, this earlier work was done using data from the 2012–2014 waves of HRS data collection). Our aims are fourfold. First, we estimate the difference in cognitive functioning between web- and phone-based respondents in 2018 based on a randomized experiment embedded into the HRS design (including attempts to account for the role of non-compliance as not all respondents assigned to the web-based mode completed the survey in that mode). Second, we attempt to use the longitudinal nature of the HRS (and the traditional usage of both phone- and in-person modalities) to provide additional context for these results regarding the web modality. Third, we examine potential variation across item types in the degree to which their functioning may vary in the web-based modality. Fourth, we attempt to probe the stability of rank-ordering of respondents across various waves in an attempt to further understand the implications of mode-based differences in survey responding, especially for widely-used cutpoints [8]. Collectively, these results are important in guiding future research that utilizes the web-based cognitive measures and also suggest possible avenues for improving the web-based measurement of cognitive functioning.

## 2 Methods

### 2.1 Data

The Health and Retirement Study is a biannual survey of US respondents over 50 and their spouses. It is designed to provide information about the economic well-being and the health of this population; more information about its design is available elsewhere [2]. We focus here on the cognitive measure collected in this survey. The HRS measure of cognitive functioning [5] is based on the TICS survey for assessing cognitive status via the telephone [9]. It includes tasks that tap respondents’ memory, mental status, and vocabulary. Although we utilize a latent variable approach, we consider many of the same data elements that are used to construct the (widely-used) cogtot measure constructed by RAND [9].

Prior to 2018, the cognitive measure were collected using phone and face-to-face modalities. Earlier work has examined whether mode effects may challenge measurement when the survey is administered via both phone and face-to-face modalities [10]; results tended to support the exchangeability of scores from those two measures. Responses would alternate from one mode to the other across consecutive waves of HRS data collection. More recent research [7] has begun to challenge the view that the modalities are interchangeable; the situation is now more complex with the introduction of the web-based mode. We focus here on an experiment embedded into the HRS’s 2018 administration that allows for reasonably straightforward analysis of mode effects; we now turn to details of this experiment.

### 2.2 Experimental Design

So as to study the potential mode effect of introducing the cognitive interview via the web, the HRS embedded an experiment. Respondents that took the 2016 survey face-to-face were randomly assigned to either the phone or web-based 2018 survey. There were a variety of eligibility requirements.^1^ In particular, eligibility was restricted to households empaneled prior to 2016 in which all respondents identified in their most recent prior interview as internet users. Households in which either respondent last completed an interview in Spanish, by proxy, or in the nursing home were excluded, as were households with a pending baseline, exit, or post-exit interview. In total, 3632 respondents were randomly assigned with 2258 selected for web and 1374 phone-based controls. After further restrictions, we focus analysis on a subset of 2740 (1052 of whom were assigned to phone and 1688 to web).^2^

One crucial challenge is that respondents assigned to web-based cognitive surveying could instead opt for phone-based surveying; we denote those who were assigned to web-based responding and completed it in that modality as compliers and those assigned to the web but who used the phone-based modality as non-compliers (N=371). If those respondents are, for example, older or have lower levels of cognitive functioning, then this might lead to bias in our estimates of the mode effect. We thus attempt to model such non-compliance in our analysis of the mode effect. These models are based on a set of covariates pertaining to respondent age and sex, cognitive functioning at prior wave^3^, self-reported health and number of chronic conditions, and whether the respondent was partnered. Descriptive statistics for these variables are shown in Table 1. Compliers and non-compliers are of similar age, but non-compliers are more likely to be female, have a lower level of educational attainment and lower cognitive functioning in 2016, have more health problems, and be unpartnered.

**Table 1:** Descriptives related to mode assignment and compliance/non-compliance

|  | N | Mean | Standardized Means |  |  |
| --- | --- | --- | --- | --- | --- |
|  |  |  | Assigned Phone | Assigned Web |  |
|  |  |  |  | Compliers | Non-compliers |
| Age | 2740 | 67.96 | 0.01 | -0.01 | 0.01 |
| Female | 2740 | 0.59 | 0.01 | -0.03 | 0.07 |
| Highest degree | 2740 | 3.10 | 0.02 | 0.01 | -0.10 |
| Cognition (2016) | 2664 | -0.01 | -0.02 | 0.06 | -0.18 |
| Self-reported health (2018)<sup>a</sup> | 2740 | 2.63 | 0.00 | -0.09 | 0.30 |
| # Chronic Conditions (2018) | 2740 | 2.26 | 0.01 | -0.07 | 0.22 |
| Partnered (2018) | 2740 | 0.64 | -0.04 | 0.13 | -0.37 |

<sup>a</sup> 1=Excellent, 5=Poor

### 2.3 Analysis

#### 2.3.1 Prediction of non-compliance

Roughly 22% of respondents assigned to the web selected out of web-based responding in favor of taking the survey via phone. We attempt to model this non-compliance using logistic regression; that is, we model

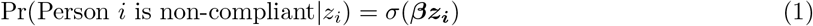

where *σ* is the logistic sigmoid 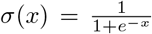 and ***z***_***i***_ is a vector of predictors. We consider three sets of predictors. The first set is based on respondent demographics. Respondent sex and race are included as well as respondent age; given that age effects may be nonlinear, we first map respondent age onto a b-spline basis [11]. The second set is respondent cognitive functioning in 2016 as computed via the IRT approach described below. The third set is a saturated model including the first two sets as well as the additional variables shown in Table 1. Probabilities generated via Eqn 1 will be used to adjust subsequent analysis to account for non-compliance as we describe below.

#### 2.3.2 IRT models

We use IRT models [6] for the purposes of modeling responses. These models suppose that the cognitive functioning of respondent *i* is captured by *θ*_*i*_ and that item functioning is captured by some set of parameters.

For simplicity (and given the primary importance of uniform forms of bias), we utilize the Rasch model such that item functioning is completely characterized by difficulty parameters. Given that the cognition data takes the form of both dichotomously and polytomously scored items, we use the partial credit model formulation ([12]; specifically see the R function gpcmIRT in [13]). For item *j*, the difficulty of the *k*-th response option is *β*_*jk*_. If item *j* is scored as 0, 1, …, *K* 1 (i.e., it has scores in *K* categories), we can write the probability of person *i* scoring in the *k*-th category as

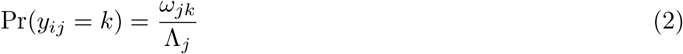

where

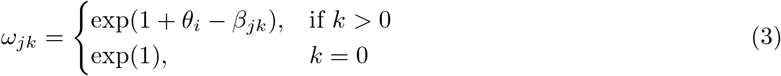

And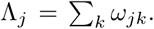. Note that if *k* = 2 Eqn 2 simplifies to the standard one-parameter logistic model for dichotomous item responses:

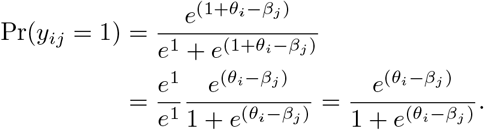

For those unfamiliar with IRT models, note that this model has a form similar to that of common logistic regression models; the challenge in this setting is that neither *θ*_*i*_ or *β*_*j*_ is directly observed.

Suppose we have two groups, *A* and *B* (i.e., phone- and web-based survey modality). Multiple-group models [14] allow us to assume separate priors on ability for respondents of different groups. We will assume that 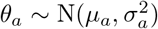 and 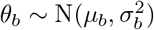. For purposes of identification, we’ll assume that *µ*_*a*_ = 0 and 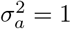. We will then estimate, using the R function multipleGroup in [13], *µ*_*b*_ and 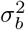 which will be the basis for our identification of the overall mode effect. Code to replicate analysis is available.^4^

#### 2.3.3 Adjustment for non-compliance

Conventional analysis of experiments would use the propensity weights in, for example, inverse probability weighting schemes to correct for non-compliance [15]. The IRT-based approach we utilize here for identification of the treatment effect does not readily allow for such weighting, so we consider two alternative approaches using the propensity weights. We first exclude phone-based respondents whose propensity score (calculated via Eqn 1) is above the *τ*-th quantile of the distribution of those who selected out of web-based responding. Such an approach is primarily intended to be suggestive since they do not clearly lead to groups that can be compared (e.g., it is not simply the web-based respondents who were above a certain quantile on the propensity score that selected for phone-based responding). We next simulated 10 datasets where inclusion for respondents assigned to phone was randomly decided via the propensity weights (e.g., for a weight of *w*, a respondent was in the sample with probability Bernoulli(1 − *w*)); we then separately analyze these data and take the mean of *µ*_*b*_ and 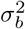 across these 10 simulated datasets.

## 3 Results

### 3.1 Understanding non-compliance

We first focus on the problem of non-compliance (i.e., the fact that people assigned to the web-based modality could opt to take the phone survey). We quantify our ability to predict non-compliance using the AUC [16] and the IMV [17]. As we may anticipate given results in Table 1, demographics (i.e., age (splined), gender and race (as a factor)) and education (highest degree) are clear predictors of non-compliance (AUC=0.64). Their strength can be compared to various benchmarks. For example, the AUC is far less than AUCs of above 0.8 for predicting mortality in these data [18] using similar demographic information.

We also consider cognition in the 2016 wave as a predictor. As a stand-alone predictor, cognition in the prior wave is a relatively weak predictor of non-compliance (AUC=0.57). The fact that cognition is a weak predictor is reassuring given that it suggests non-compliance might be largely ignorable for the purposes of estimating the treatment effect. We then consider models that include all variables from Table 1. Crucially, cognition in 2016 does not provide additional information net of these other predictors and also leads to some missingness (the IMV calculated via out-of-sample data suggests that inclusion of the cognition variable results in only very minor changes to prediction quality). Thus, we use predictions from the full model minus cognition (final row in Table 2) in subsequent analysis that utilize propensity weights.

**Table 2:** Predicting non-compliance

| Predictors | N | N non-compliant | AUC | IMV |
| --- | --- | --- | --- | --- |
| Demo+Edu | 1688 | 371 | 0.642 | 0.016 |
| Cog (2016) | 1624 | 358 | 0.572 | 0.005 |
| Full<sup>a</sup> | 1624 | 358 | 0.703 | 0.035 |
| Full (no Cog) | 1688 | 371 | 0.705 | 0.032 |

<sup>a</sup> All variables in Table 1.

### 3.2 Overall mode effects

Using multiple-group IRT estimation, we can assess overall mode effects. We first do so via unadjusted comparison of respondents who completed the cognitive survey according to their assigned modality (i.e., ignoring non-compliance); see the top row of Table 3. This analysis suggests a relatively large mode effect of nearly 1/3 of a standard deviation of respondent ability on the phone-based survey, a potentially serious problem especially given that earlier work on mode effects related to phone/in-person differences found negligible results [5]. However, this estimate may be biased given that it does not adjust for non-compliance.

**Table 3:** Estimated group differences

| | N phone | N web | $\mu_{\text{web}}$ | $\sigma_{\text{web}}^2$ |
| --- | --- | --- | --- | --- |
| Unadjusted | 1052 | 1317 | 0.32 | 0.62 |
| $\tau_{0.99}$ | 1049 | 1317 | 0.32 | 0.62 |
| $\tau_{0.95}$ | 1024 | 1317 | 0.30 | 0.62 |
| $\tau_{0.9}$ | 1000 | 1317 | 0.28 | 0.62 |
| $\tau_{0.8}$ | 942 | 1317 | 0.26 | 0.62 |
| Random | 809.80 | 1317.00 | 0.29 | 0.60 |

We consider two ways of adjusting for non-compliance (see Section 2.3.3). We first restrict ourselves to data with respondents who are relatively unlikely to be non-compliant (based on estimates from Eqn 1). We then use estimates from Eqn 1 to generate multiple datasets in an attempt to simulate the process of compliance. Both approaches—which lead to estimates of approximately 0.29 to 0.32—suggest that selection (as modeled by the given predictors) leads to only a slight upward bias in the observed mode effect. Given the relatively mild impact of non-compliance, we do not adjust for it in the analyses below; while this might lead to some bias in the resulting estimates it also allows for more straightforward analysis.

### 3.3 Longitudinal Analysis

We can use the biennial nature of the HRS to get a longitudinal view of the issue as well. Figure 1 (Panel A) takes the multigroup IRT estimation strategy to estimate abilities for 2161 individuals assessed between 2012 and 2018. Focusing first on the gray line, two facts are important. First, when we consider same-mode measures, respondents decline over time (scores in 2012 are higher than in 2016 and 2014 scores are higher than 2018). This is reassuring given that such a finding would be expected given the general decline in cognition as respondents age [1]. Second, there seems to be a mode effect such that phone-based scores are higher than face-to-face (f2f) scores. Turning to the red dot which shows the estimated cognitive functioning of web-based respondents, we can see that web-based respondents appear to have much higher levels of cognitive functioning. This is, of course, not plausible given the experimental design. Further, the size of the effect is much larger than the relative advantage that phone-based respondents have relative to in-person respondents in the previously used modalities.

**Figure 1:**
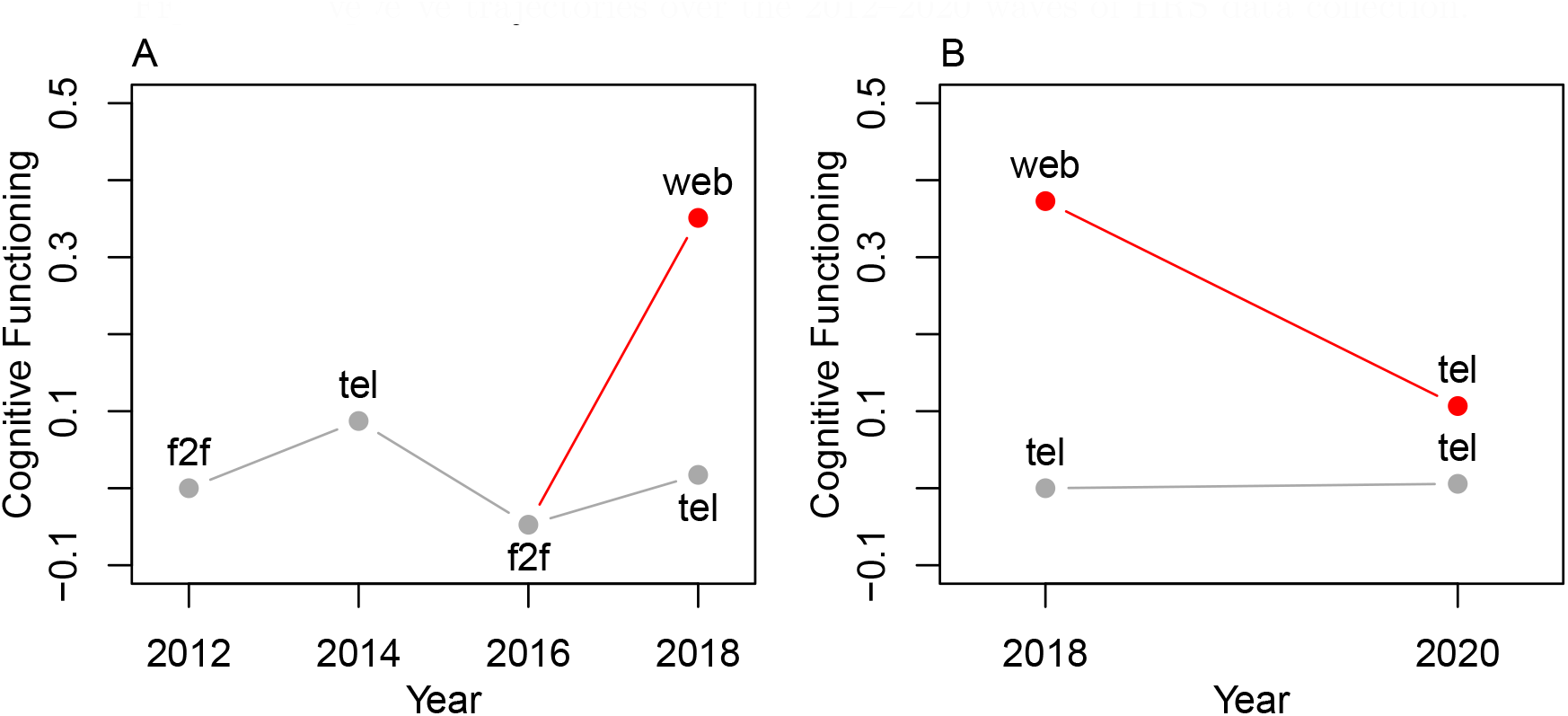
Cognitive trajectories over the 2012–2020 waves of HRS data collection.

We can also use the fact that respondents from this experiment largely took the phone-based cognitive survey in the 2020 wave of data collection. Figure 1 (Panel B) shows that, of the respondents in the experiment who also were assessed in 2020, we see a substantial decrease in functioning for the web-based group in 2018 when they are assessed via phone in 2020. In contrast, the phone-based group performs similarly at both waves.

### 3.4 Item-specific mode effects

We conducted item-specific analysis to test whether mode effects are homogeneous or vary in severity across items. Given the nature of the HRS cognitive interview, we considered four bundles of items: the delayed recall items, the immediate recall items, the serial 7s, and the financial items. Results are in Table 4. We first note the standardized difference on the standardized sum score across the bundle. Note that web respondents score higher across all bundles; if all bundles show some degree of bias towards web-based respondents, this makes it challenging to identify differential item functioning across any given bundle in an unbiased manner [19].

**Table 4:** Item-specific mode effects

| | $\Delta_{\text{standardized}}^a$ | DIF | | Multigroup | |
| --- | --- | --- | --- | --- | --- |
| | | Estimate | Std. Error | $\mu_{\text{web}}$ | $\sigma_{\text{web}}^2$ |
| Delayed | 0.19 | -0.14 | 0.02 | 0.42 | 0.44 |
| Immediate | 0.19 | -0.14 | 0.02 | 0.42 | 0.41 |
| Serial 7 | 0.38 | 0.23 | 0.03 | 0.37 | 0.91 |
| Finances | 0.53 | 0.39 | 0.03 | 0.31 | 0.91 |

<sup>a</sup> $z_{\text{web}} - z_{\text{tel}}$ when $z$ has been standardized.

That said, we can still assess differences in relative magnitudes. We conduct basic differential item functioning (DIF; [20]) analyses wherein we regress (using OLS) the standardized bundle score on the overall mean score plus a group indicator. In the DIF columns, we focus on the coefficient associated with web-based responding. Note that the recall items (both delayed and immediate) show negative coefficients. This is due to the problem mentioned above [19] and should be interpreted as suggesting that bias in favor of web-based responding is less severe on these items as compared to the Serial 7s and Finance items. In the multigroup columns, we re-estimated group mean differences after dropping each bundle. These analyses confirm that the bias is most severe on the Serial 7 and the Finance items with the Finance items showing the most severe bias across all analyses.

### 3.5 Stability of respondent rank ordering

The above results suggest that the web-based version of the cognitive assessment was easier than the phone-based version. This has implications for the use of this data which is a point we return to in the discussion. A different question is whether the test was differentially easier for some respondents as compared to others such that it changed the relative ranking of respondents. If the web-based version is just uniformly easier, then it might still be possible to establish cutpoints (e.g., [8]).

To analyze this issue, we consider the rank-order correlations (i.e., Spearman correlations) of 2016 cognitive scores (specifically the cogtot score) with 2018 estimates (via the IRT approach emphasized here) separately across response modality. Results are in Figure 2. The overall stability is fairly comparable across modes with the estimates for phone-based respondents having a Spearman correlation of 0.52 with the 2016 total cognition scores whereas the web-based scores are correlated with 2016 totals at 0.46. We also stratified rank-order correlations by age; see Table 5. While there is clear variation, the results are generally consistent with the findings from Figure 2 wherein phone-based correlations are somewhat higher than web-based estimates.

**Table 5:** Rank-order correlations by age bins

| Age | N | Tel | Web |
| --- | --- | --- | --- |
| All | 2740 | 0.52 | 0.46 |
| (50,60] | 529 | 0.22 | 0.61 |
| (60,70] | 1205 | 0.49 | 0.43 |
| (70,80] | 683 | 0.53 | 0.46 |
| (80,Inf] | 281 | 0.46 | 0.36 |

**Figure 2:**
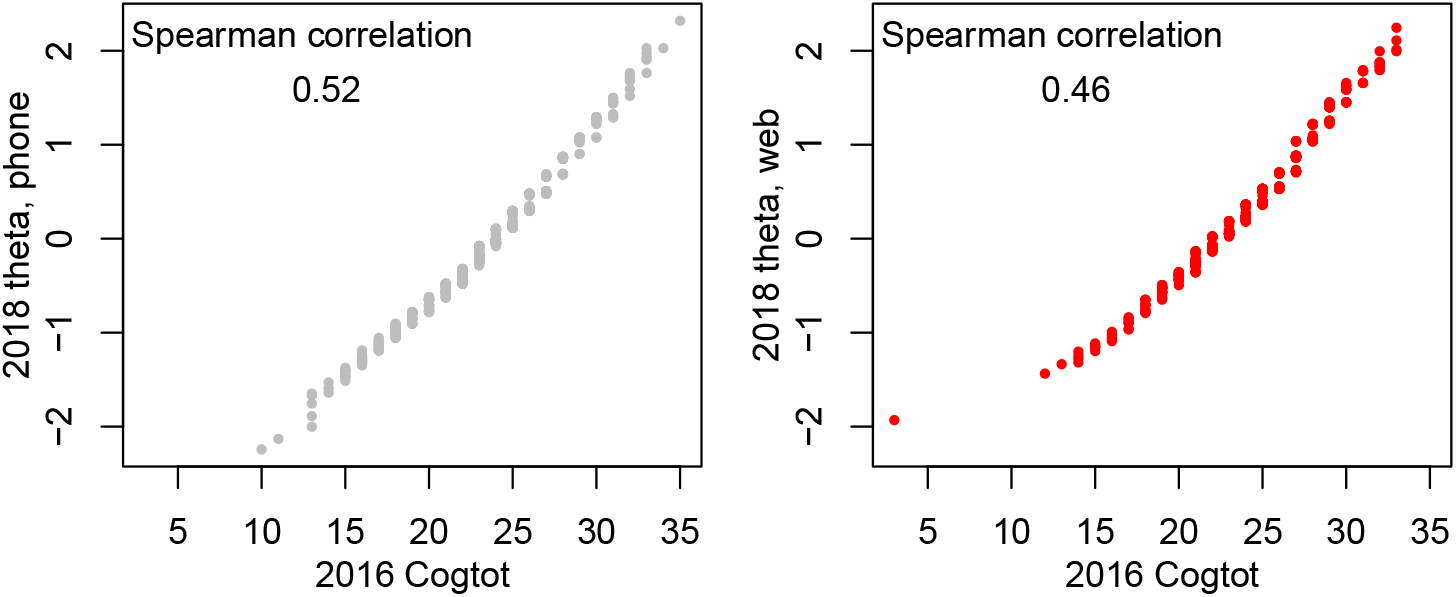
Stability of rank orders

### 3.6 Implications for Langa-Weir Cutpoints

We finally consider the implications for the widely-used Langa-Weir cutpoints [8]. These cutpoints are based on the 27-point sum score scale defined by the recall, backward counting, and sum score items. We first discuss the left panel of Figure 3. This panel is based on aggregated *θ* scores computed via multigroup IRT. For the phone-based respondents, we use their observed score on the 27 point scale underlying the Langa-Weir classifications [21]. The x-axis shows the average *θ* score for each of the different points. For the web-based respondents, we imputed scores (using previously established techniques [22]). These results suggest a fairly consistent increase in web-based *θ* values relative to phone-based *θ* values for a similar sum score. Note that this analysis relies on two assumptions. First, we use imputed scores for web-based respondents. This assumption relies upon the item-specific mode effects (as per Section 3.4) for the unobserved items being consistent with those observed here. Second, we ignore attrition here.

**Figure 3:**
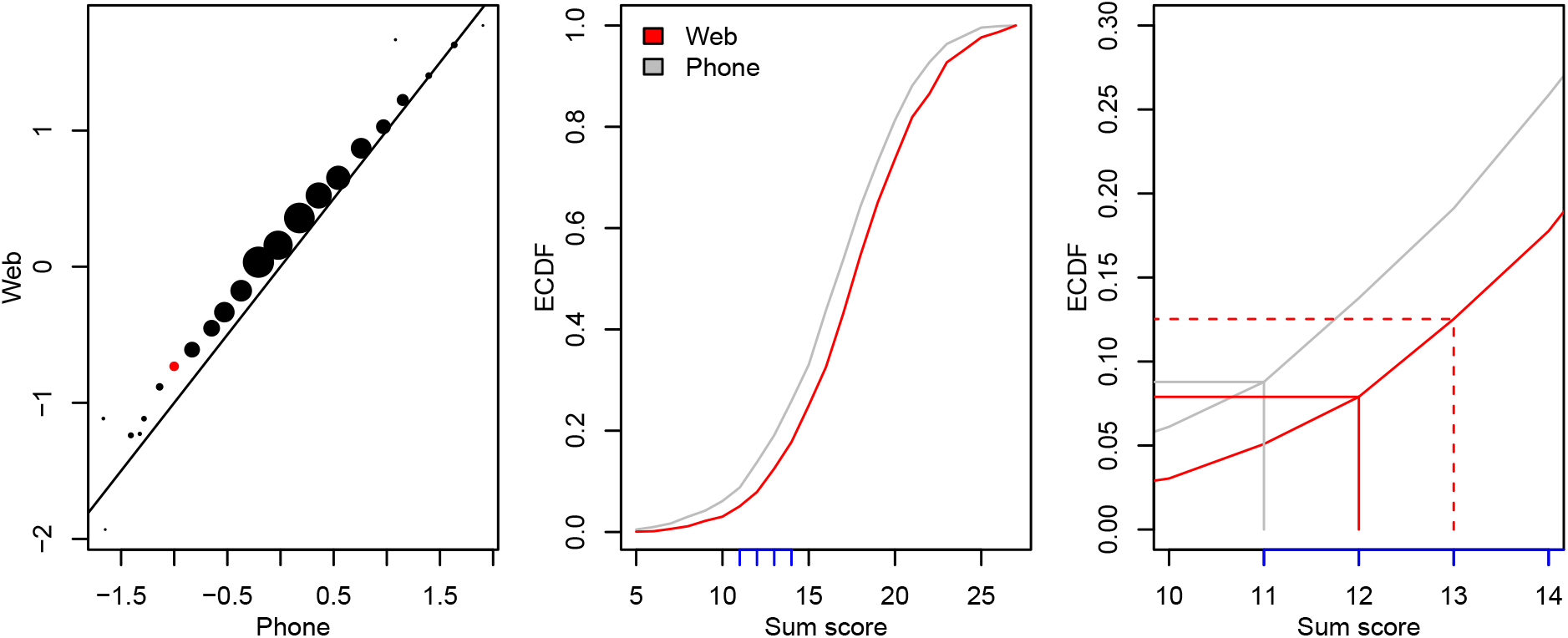
Analysis of scores related to Langa-Weir cutpoints [8]. Left: Scatterplot of mean IRT scale scores for phone (actual) and web (imputed) respondents with dots scaled to represent the amount of data for each score. Middle: ECDF for web- and phone-based sum scores. Right: Cutpoint analysis based on ECDFs

In the middle panel of Figure 3, we compute empirical cumulative density functions (ECDF) for the sum scores used in the left panel. Note that the web-based scores are uniformly right-shifted relative to the phone-based scores which is consistent with the notion of a relative constant mode effect. Of particular interest are the points on the x-axis shown in blue which pertain to a key cutpoint in the Langa-Weir framework [8]. In the right panel, we focus on the implications for the threshold separating those respondents who are cognitively impaired but not demented (CIND) from those exhibiting normal functioning. On the 27-point scale, it is a value of 11 when assessed using conventional modalities. Here, 8.8% of the phone-based respondents are at or below below that cutscore (note the segments in gray). Given the random equivalence of the web- and phone-based groups, we look for the sum score at which similar proportions of web-based respondents are less than or equal to. Of the web-based respondents, 7.9% get less than or equal to a 12 (12.5% get less than or equal to a 13; see dashed red line). Thus, we argue that a cutscore of 12 would be superior to a cutscore of 11 as the maximal score for a person to be classified as CIND.

## 4 Discussion

Mode effects can be a serious threat to the inferences made based on psychological measures. Given the ubiquity of devices and the relative challenge of getting people to respond to surveys via the phone, a transition to web-based surveying is of clear interest to surveys like the HRS. However, that transition also leads to challenges associated with the possibility of mode effects. Using the experiment embedded in the 2018 survey, we find that web-based respondents do much better than expected on the survey; these results align with others [23] suggesting some degree of difference in web-based responding on the HRS surveys. While we do not test hypotheses for why this might be, one clear possibility is that respondents are making use of the affordances offered by a computer (e.g., computational power) in a way that they do not when responding via phone. We focus on comparisons to phone-based assessment, but recent reports [24] (see also Figure 1 here) suggest that phone-based assessment may itself have a mode effect such that it is easier than in-person assessment which would further complicate comparison of web and face-to-face scores.

One potential threat to the study design is that respondents assigned to web-based responding could still opt into phone-based responding. While there was some clear patterning as to what kinds of respondents made that switch (e.g., they were less well-educated and in relatively poorer health), adjustments for this type of non-compliance suggested that it led to fairly minimal bias in our estimates of the mode effect. We also took advantage of the longitudinal structure of the HRS data to put the web effect in context. Web-based respondents both show outsized gains on the 2018 survey but the web effect is also much larger than the relatively modest advantage that accrues to phone-based responding vis-a-vis in-person responding. Further, there is a large decline in 2020 when the survey is administered via phone for those who took the survey in 2018 via the web.

We acknowledge limitations. A primary one pertains to the issue of generalizability and is based on the fact that respondents had to meet fairly specific criteria to be included in the experiment. Respondents who are not as familiar (due to, for example, age) with digital devices may not benefit in the same way from web-based responding. Further, our results may not be informative about respondents suffering from serious cognitive impairment as they also were not eligible. In addition, other approaches are available for analysis of mode effects (e.g., [25]); usage of such approach may provide additional information about the role of web-based responding in how the HRS measures cognition.

### 4.1 Implications and Recommendations

Our findings have implications for the HRS. For clarity, we list them below:

- Direct comparisons of scores derived from the web to those from other modalities will likely be misleading due to the existence of mode effects. In particular, web-based scores can be expected to be slightly higher than phone-based scores.
- Adjustments so as to make scores directly comparable are perhaps possible but will need to be used with great care.
- In studies that focus on classifications of cognitive function, classifications of CIND from the web-based measure may want to use a threshold of 12 and below (in place of 11 and below that has been used previously [21]).

In general, as more of the HRS transitions to web-based assessment, there will be a keen need for attention to the resultant changes in what the HRS is measuring about respondents. There is great wisdom in the classic maxim on this point—“If you want to measure change, don’t change the measure.” [26]—but changes in people’s lives will necessitate changes in measurement; surveys like the HRS will need to use techniques along the lines of those deployed here to help calibrate the impacts of such changes on measures of key quantities.

## Data Availability

All data in the present work are available from the HRS website: https://hrs.isr.umich.edu/data-products.

## Acknowledgements

This project was funded in part by the Jacobs Foundation.

## Footnotes

1 Here is official HRS documentation on the matter: “In 2018, for the first time HRS added Web as an alternate mode for data collection for the core biennial interview. Web was offered as an alternative for telephone and regular face-to-face respondents only; respondents in the half-sample scheduled for the enhanced face-to-face interview were not eligible for Web this wave. Among the half-sample scheduled for telephone or regular face-toface in 2018, there were just over 3,700 sample members who were eligible for Web based on a prior report of internet access along with other selection criteria (e.g., English speaking, self-respondent, non nursing-home resident in prior wave). Of the 3,700 Web eligible sample members, 60% were randomly selected for the Web sample with the remainder receiving their usual mode of either telephone or face-to-face for comparison purposes. Participants in the Web sample received an advance letter inviting them to complete their 2018 interview on the Web and containing a URL to access the survey. This was followed two days later by an email with a link to the survey. A phone number was provided for respondents to contact us if they preferred to complete their interview by phone or needed assistance with the Web mode. Email and mail reminders were sent on a regular schedule. Participants in the Web sample who had not completed the survey by Web after a predetermined period of no activity were contacted for a telephone interview and were able to resume the survey where they left off in the new mode. The Web survey was programmed to be as close as possible to the other modes. Rather than break the survey into components we designed it to allow respondents to work at their own pace and to stop at any point and return later to continue, even if they required several sessions to complete it.” https://hrsdata.isr.umich.edu/sites/default/files/documentation/data-descriptions/1652380440/h18dd.pdf

2 We excluded 143 respondents who responded via one of our non-focal modalities (e.g., face-to-face or via the small web module) and 749 non-respondents.

3 Cognitive functioning at the prior wave was calculated using the multigroup IRT approach introduced in Section 2.3.2.

4 https://github.com/ben-domingue/hrsweb

